# Direct Measurement of Rates of Asymptomatic Infection and Clinical Care-Seeking for Seasonal Coronavirus

**DOI:** 10.1101/2020.01.30.20019612

**Authors:** Jeffrey Shaman, Marta Galanti

**Affiliations:** Department of Environmental Health Sciences, Mailman School of Public Health, Columbia University, 722 West 168th Street, New York, NY 10032

## Abstract

The pandemic potential of the novel coronavirus (nCoV) that emerged in Wuhan, China, during December 2019 is strongly tied to the number and contagiousness of undocumented human infections. Here we present findings from a proactive longitudinal sampling study of acute viral respiratory infections that documents rates of asymptomatic infection and clinical care seeking for seasonal coronavirus. We find that the majority of infections are asymptomatic by most symptom definitions and that only 4% of individuals experiencing a seasonal coronavirus infection episode sought medical care for their symptoms. These numbers indicate that a very high percentage of seasonal coronavirus infections are undocumented and provide a reference for understanding the spread of the emergent nCoV.

## Background

The Wuhan novel coronavirus (nCoV) appears to have emerged in humans in the Hubei province of China during December 2019 (*1*). The virus was definitively identified in the first week of 2020 and as of January 29 had spread to 15 other countries; 6065 confirmed cases, 9239 suspected cases, and 132 deaths have been reported (*2*); however, the total number of infections (confirmed, suspected and unreported) remains unknown.

Quantification of the reporting rate, i.e. the ratio of confirmed and suspected cases to total infections, is vital for delineation of the prevalence, scope and potential spread of nCoV. Indeed, the speed and geographic extent of its translocation throughout China and internationally are likely tied to the true prevalence of the pathogen and suggest that there are many more infections (i.e. the reporting rate is low), that contagiousness is not negligible among these undetected infected persons, and that nCoV will be difficult to contain. To contextualize the issue of unreported infection, we here present the findings from a recent proactive sampling project carried out in New York City (NYC) that documented rates of asymptomatic infection and clinical care-seeking among individuals shedding seasonal CoV (types: HKU1, 229E, NL63 and OC43).

## Methods

Data are derived from sampling performed between October 2016 and April 2018 as part of the Virome project, a proactive sampling of respiratory virus infection rates, associated symptom reports and rates of seeking clinical care. We enrolled 214 healthy individuals from multiple locations in the Manhattan borough of NYC. Cohort composition is described in Galanti et al. (*3,4*) and includes: children attending two daycares, along with their siblings and parents; teenagers and teachers from a high school; adults working at two emergency departments (a pediatric and an adult hospital); and adults working at a university medical center. The cohort was obtained using convenience sampling, and all participants were younger than 65 years. While the study period spanned two years from October 2016 to April 2018, some individuals enrolled for a single cold and flu season (October – April) and others for the entire study period. Participants (or their guardians, if minors) provided informed consent after reading a detailed description of the study (CUMC IRB AAAQ4358).

Nasopharyngeal samples were collected by study coordinators once a week irrespective of participant symptoms. Samples were screened using the GenMark eSensor RVP system for 18 different respiratory viruses, including coronavirus 229E, NL63, OC43, and HKU1. Sample collection and extraction followed the same protocol as in Shaman et al. (*5*).

In addition, participants completed daily self-reports rating nine respiratory illness-related symptoms (fever, chills, muscle pain, watery eyes, runny nose, sneezing, sore throat, cough, chest pain) each of which was recorded on a Likert scale (0=none, 1=mild, 2=moderate, 3=severe). The daily self-report also documented whether participants had sought medical attention, stayed home or taken cold and flu-related medications (both over-the-counter and prescription) as a consequence of their listed symptoms (see (*4*) for further survey details).

We defined an infection (or viral) episode as a group of consecutive weekly specimens from a given individual that were positive for the same virus (allowing for a one-week gap to account for false negatives and temporary low shedding). Medically attended illness, sick days at home and medicine uptake were defined as episodes in which the participants reported seeking care, staying home or taking medicines due to respiratory symptoms. We classified all infection episodes as symptomatic or asymptomatic according to individual symptom scores in the days surrounding the date of the first positive swab of an episode. We used multiple definitions as a standard for symptomatic infection does not exist (Table 1). These symptom definitions are described in reference to a −3 to +7-day window around the date of the initial positive swab for an infection episode. Daily score is defined as the sum of the 9 individual symptoms (range: 0-27). Total symptom score is the daily symptom score summed over the −3 to +7-day window.

**Table 1.** Definitions of symptomatic infections. All symptom definitions are described in reference to a −3 to +7-day window around the date of the initial positive swab for an infection episode. Note, Definition 4 is relative to an individual’s long-term average total symptom score.

|  |  |
| --- | --- |
| Definition 1 | At least one day with a daily score >3 |
| Definition 2 | Minimum two individual symptoms >0 and at least one symptom >1 |
| Definition 3 | Total symptom score >9 |
| Definition 4 | Total symptom score greater than twice the weekly average for the infected individual |
| Definition 5 | Total symptom score >0 (i.e. any reported symptom) |

## Results

Table 2 presents the findings for seasonal CoV from this proactive sampling effort. 135 CoV infection episodes were identified. CoV-OC43 infection was most frequent (n=61), and CoV-NL63 infection was least frequent (n=15). Note that co-infections occurred; hence, the sum of the type infections is greater than the total reported in the final column of Table 2. Adults (20+ years old) constituted 64% of the cohort, provided 67% of collected swab specimens, and experienced 50% of all CoV infection episodes.

**Table 2.** Coronavirus Infections. Estimates are obtained from the daily symptom diaries and weekly nasopharyngeal swab samples from October 2016 through April 2018.

|  |  | 229E | OC43 | HKU1 | NL63 | All |
| --- | --- | --- | --- | --- | --- | --- |
| **Infections** | All Episodes (n=214 participants; 4215 swabs) | 33 | 61 | 30 | 15 | 135 |
|  | Among Children (<20 yrs, n=35; 1016 swabs) | 11 (33%) | 32 (52%) | 14 (46.5%) | 4 (26.5%) | 57 (42%) |
|  | Among Teens (11-19 yrs; n=42; 361 swabs) | 4 (12%) | 4 (7%) | 2 (7%) | 1 (7%) | 11 (8%) |
|  | Among Adults (20+ yrs; n=137; 2838 swabs) | 18 (55%) | 25 (41%) | 14 (46.5%) | 10 (66.5%) | 67 (50%) |
| **Symptoms** | Definition 1 | 5 (15%) | 16 (26%) | 11 (37%) | 9 (60%) | 40 (30%) |
|  | Definition 2 | 9 (27%) | 13 (21%) | 8 (27%) | 9 (60%) | 39 (29%) |
|  | Definition 3 | 7 (21%) | 20 (33%) | 15 (50%) | 8 (53%) | 49 (36%) |
|  | Definition 4 | 5 (15%) | 20 (33%) | 10 (33%) | 7 (47%) | 41 (30%) |
|  | Definition 5 | 18 (55%) | 33 (54%) | 18 (60%) | 11 (73%) | 79 (59%) |
| **Care** | Medicine Uptake | 4(12%) | 16 (26%) | 10 (33%) | 6(40%) | 35(26%) |
|  | Stayed Home | 0 (0%) | 3 (5%) | 7 (23%) | 3(20%) | 13 (10%) |
|  | Sought Medical Care | 0 (0%) | 3 (5%) | 3 (10%) | 0 (0%) | 6 (4%) |

Based on Definitions 1-4, the majority of CoV infection episodes were asymptomatic. When stratified by type, differences emerge: CoV-NL63 was more than twice as likely to produce symptomatic infection episodes as CoV-229E (by Definitions 1-4). Definition 5, which defines symptomatic infection as the manifestation of any symptom during the 11-day window spanning the initial positive swab collection, indicates that a majority of infected persons reported at least one mild cold-related symptom.

Participants reported taking medicine in response to cold and flu symptoms for 26% (n=35) of CoV infection episodes. Participants were three times more likely to take medicine in association with CoV-NL63 infection (40%) than CoV-229E infection (12%). 10% of participants reported staying home from work or school during CoV infection episodes, and 4% of participants reported seeking medical care.

## Discussion

Reporting of seasonal CoV prevalence has been limited to date. The US CDC only began releasing CoV positivity rates in 2018, and understanding of its epidemiological features is incomplete. Here, we present direct measurement of rates of seeking clinical care and show that only 4% (6 of 135) of persons shedding seasonal CoV sought care. Reporting rates and rates of seeking medical care may differ, as most individuals visiting a clinician for acute respiratory illness are not tested for seasonal CoV. However, testing for the 2019 nCoV is expected to be more robust than for seasonal CoV as there is pressing need to understand and contain this novel pathogen. This high testing rate could result in the reporting rate for nCoV equaling the rate of seeking medical care. Thus, if rates of seeking medical care for nCoV were to align with seasonal CoV, the reporting rate for nCoV would approach 4%. Should the remaining 96% of unreported infections be sufficiently contagious, their high numbers could support rapid spread of nCoV.

It is important to note that rates of seeking medical care for nCoV may not align with the rates found here for seasonal CoV: concern over the virus may lead to elevated rates of seeking medical care; our estimates for seasonal CoV represent a moderate sample size; and the symptom severity of emergent Wuhan nCoV may differ from seasonal CoV. Further, early outbreak instances of death due to nCoV occurred predominantly in elderly patients, and often elderly with co-morbidities (*6*). The data we present here for seasonal CoV are from a cohort with no individuals 65 year or older, so the findings identified here might not have bearing on the elderly.

Still, several recent studies suggest that reporting rates for nCoV may align with the rates found here for seasonal CoV. Chinazzi et al., (*7*) used an agent-based modeling approach to estimate the overall nCoV incidence (reported and unreported) necessary to support the observed numbers of geographically exported infected individuals as of January 21, 2020. They estimated a 5-16% nCoV reporting rate depending on model assumptions. In addition, phylogenetic analysis has identified Wuhan nCoV as a betacoronavirus, the genus that includes CoV-OC43 and CoV-HKU1, as well as CoV-SARS and CoV-MERS (*8*). Among our cohort participants, 7% (6 of 91) of infection episodes with seasonal betacoronavirus (OC43 and HKU1) sought medical care.

The impact of unrecognized and unattended infections on transmission dynamics is likely a key determinant of whether control of the nCoV outbreak is possible. For example, although cases of seroconversion without sign of disease were documented during the SARS outbreak, asymptomatic and subclinical rates are believed to have been low. No cases of asymptomatic SARS transmission were documented (*9*), which may have facilitated the containment of the epidemic.

A higher asymptomatic ratio was reported for MERS infections, which presented with a broader spectrum of clinical manifestations: 21% of laboratory identified cases were mild or asymptomatic and 48% had severe disease or died (*10*). The potential for asymptomatic transmission of MERS has been reported (*11*); however, the majority of transmission events happened within hospital or household, suggesting that close proximity may have facilitated transmission.

nCoV has spread rapidly in a short time, which suggests a substantial number of unreported infections exist. The results presented here provide a unique direct measure of seasonal CoV infection, symptom severity, and rates of seeking medical care. These findings provide a baseline reference for contextualizing nCoV pandemic potential.

## Data Availability

All data are available upon request.

